# Patterns of Multimorbidity and Risk of Severe SARS-CoV-2 Infection: an observational study in the U.K

**DOI:** 10.1101/2020.10.21.20216721

**Authors:** Yogini V Chudasama, Francesco Zaccardi, Clare L Gillies, Cameron Razieh, Thomas Yates, David E Kloecker, Alex V Rowlands, Melanie J Davies, Nazrul Islam, Samuel Seidu, Nita G Forouhi, Kamlesh Khunti

## Abstract

**Background:** Pre-existing comorbidities have been linked to SARS-CoV-2 infection but evidence is sparse on the importance and pattern of multimorbidity (2 or more conditions) and severity of infection indicated by hospitalisation or mortality. We aimed to use a multimorbidity index developed specifically for COVID-19 to investigate the association between multimorbidity and risk of severe SARS-CoV-2 infection.

**Methods:** We used data from the UK Biobank linked to laboratory confirmed test results for SARS-CoV-2 infection and mortality data from Public Health England between March 16 and July 26, 2020. By reviewing the current literature on COVID-19 we derived a multimorbidity index including: 1) angina; 2) asthma; 3) atrial fibrillation; 4) cancer; 5) chronic kidney disease; 6) chronic obstructive pulmonary disease; 7) diabetes mellitus; 8) heart failure; 9) hypertension; 10) myocardial infarction; 11) peripheral vascular disease; 12) stroke. Adjusted logistic regression models were used to assess the association between multimorbidity and risk of severe SARS-CoV-2 infection (hospitalisation or death). Potential effect modifiers of the association were assessed: age, sex, ethnicity, deprivation, smoking status, body mass index, air pollution, 25‐hydroxyvitamin D, cardiorespiratory fitness, high sensitivity C-reactive protein.

**Results:** Among 360,283 participants, the median age was 68 [range, 48-85] years, most were White (94.5%), and 1,706 had severe SARS-CoV-2 infection. The prevalence of multimorbidity was more than double in those with severe SARS-CoV-2 infection (25%) compared to those without (11%), and clusters of several multimorbidities were more common in those with severe SARS-CoV-2 infection. The most common clusters with severe SARS-CoV-2 infection were stroke with hypertension (79% of those with stroke had hypertension); diabetes and hypertension (72%); and chronic kidney disease and hypertension (68%). Multimorbidity was independently associated with a greater risk of severe SARS-CoV-2 infection (adjusted odds ratio 1.91 [95% confidence interval 1.70, 2.15] compared to no multimorbidity). The risk remained consistent across potential effect modifiers, except for greater risk among men.

**Conclusion:** The risk of severe SARS-CoV-2 infection is higher in individuals with multimorbidity, indicating the need to target research and resources in people with SARS-CoV-2 infection and multimorbidity.

## INTRODUCTION

The novel coronavirus SARS-CoV-2 has led to a global pandemic of a complex, pneumonia-related illness (COVID-19). As of 29^th^ September 2020, there have been 33,749,599 confirmed cases of COVID-19 and 1,009,829 deaths reported worldwide.[1] Many countries are still facing a devastating rise in the number of cases and deaths, whist some are facing a second wave such as France, Spain, UK, with significant impacts on their healthcare systems. The risk of SARS-CoV-2 infection is based on the person-to-person transmission rate at which the virus is circulating in the community.[2, 3] Yet, while some individuals recover from the SARS-CoV-2 infection, others develop severe illness which may require intensive care in hospital or result in mortality.

Several potential factors have been postulated or associated with the increased risk of transmission and development of severe SARS-CoV-2 infection, such as age, ethnicity, body mass index (BMI), smoking, deprivation, pre-existing comorbidities (particularly diabetes, hypertension and cardiovascular disease), cardiorespiratory fitness, 25‐ hydroxyvitamin D level, air pollution and inflammation.[4] However, individuals with pre-existing health conditions have been found to have the highest risk of developing severe SARS-CoV-2 infection, as shown in a number of studies from early research based in Wuhan [5] to recent studies across the globe.[6-8] Nevertheless, the majority of research to date has focused on pre-existing single chronic conditions,[9] yet individuals often have multiple chronic conditions, known as multimorbidity. In view of the rapidly increasing prevalence of severe SARS-CoV-2 infection across the globe, clarifying the pattern and impact of multimorbidity (defined as 2 or more conditions) on the risk of severe SARS-CoV-2 infection has important clinical and public health implications which are not captured by a focus on single diseases.[10]

There are a number of methods to define multimorbidity, which include both simple measures such as summing up the diseases and more complex measures using a weighted score such as the Charles Comorbidity Index. A previous study in UK Biobank examined the pattern of the most prevalent clusters of multimorbidity in those infected with SARS-CoV-2, using 36 different conditions to define multimorbidity,[11] though this masked the most relevant conditions related to SARS-CoV-2.[12] Moreover, three studies which assessed the risk of multimorbidity and COVID-19 had also used a broad range of conditions (ranging from 19 to 43 conditions).[13-15] In contrast, the available evidence indicates that certain health conditions are associated with a much greater risk of severe SARS-CoV-2 infection linked with hospitalisation or mortality.[5, 16]

To help clarify the evidence, in this study we aimed to firstly develop a multimorbidity index of the most commonly reported chronic conditions from the current literature for severe SARS-CoV-2 infection. The index was then used to investigate the patterns and association between multimorbidity and risk of severe SARS-CoV-2 infection and we further tested whether this association was modified by other potential risk factors.

## METHODS

This study is reported as per the Strengthening the Reporting of Observational Studies in Epidemiology (STROBE) guidelines (***Additional file: Checklist S1***) and following a pre-specified protocol (Application Number 36371).[17]

### Study population

UK Biobank included half a million middle-aged adults recruited from 22 sites across England, Wales and Scotland with baseline measures collected between 2006 and 2010, from an in-person baseline interview at the UK Biobank centre (http://biobank.ctsu.ox.ac.uk/crystal/search.cgi). Written informed consent was obtained prior to data collection; UK Biobank was approved by the NHS National Research Ethics Service (16/NW/0274; ethics approval for UK Biobank studies).[18]

SARS-CoV-2 laboratory confirmed test results and death data for all-cause mortality from Public Health England were linked to the UK Biobank database.[19] Data were available between March 16 and July 26, 2020, restricted to those tested in a hospital (pillar 1), since this can be regarded as a proxy for hospitalisations for severe cases of the disease as suggested by the linkage methodology.[19] Our analytical sample included study participants from England as testing was available and linked, we therefore excluded study members from Scotland and Wales, participants who had an outpatient test positive for COVID-19 (pillar 2) as the outcome for SARS-CoV-2 infection was uncertain (i.e. recovered, hospitalised at a later stage, or died), those who died before March 16, 2020, and those with missing data (***Additional file: Figure S1***).

### Multimorbidity Index

To use prior evidence on comorbidities that cluster together and that may act in combination to affect the risk of COVID-19, we carried out a literature search to identify the most common pre-existing comorbidities in patients with severe SARS-CoV-2 infection (***Additional file: Table S1***). The studies from the literature search were found to be very heterogeneous, therefore it was not possible to do a quantitative meta-analysis of pooled results.[5-8, 20-29] From a semi-quantitative perspective, we selected the most common conditions to create a multimorbidity index for severe COVID-19 including the following conditions: 1) angina; 2) asthma; 3) atrial fibrillation; 4) cancer; 5) chronic kidney disease (CKD); 6) chronic obstructive pulmonary disease (COPD); 7) diabetes mellitus; 8) heart failure; 9) hypertension; 10) myocardial infarction; 11) peripheral vascular disease; and 12) stroke. Data on these comorbidities were available in UK Biobank together with the clinical marker of CKD, the estimated Glomerular Filtration Rate (eGFR) value less than 60 mL/min/1.73m^2^. Those with two or more of these conditions clustering together were assigned as having multimorbidity and the remainder without multimorbidity.

### Outcome measure

The outcome was severe SARS-CoV-2 infection, defined as admission to hospital with a positive SARS-CoV-2 infection test result or death occurring within the study period of March 16 to July 26 July, 2020. The comparison group without severe SARS-CoV-2 infection included participants with negative test result for SARS-CoV-2 either in a hospital setting or as outpatient, or having not been tested. Participants who received a positive test outside of a hospital setting were excluded from this analysis, as their outcome was uncertain (i.e. recovered, hospitalised at a later stage, or died).

### Effect modifiers

The association between multimorbidity and severe SARS-CoV-2 infection was assessed across levels of prespecified potential effect modifiers based on the literature, including: 1) age at test (grouped as <60 years or ≥ 60 years; 2) sex; 3) ethnicity (self-reported and grouped as white or non-white); 4) Townsend deprivation index (used to measure socio-economic status, which combines census data on housing, employment, and social class based on the postal code of participants; a higher Townsend score equates to higher levels of socioeconomic deprivation and participants were split in two groups based on the sample median: least or most deprived); 5) body mass index (BMI), following the exclusion of individuals in the underweight [<18.5 kg/m^2^] group (n=2,093), since <1% of the participants were categorised as underweight; the remaining participants were grouped as: normal weight [18.5-24.9 kg/m^2^], overweight [25-29.9 kg/m^2^], or obese [≥30 kg/m^2^]); 6) smoking (current smoker, previous smoker or never smoked at the time of assessment); 7) the annual average air pollution, measured by the concentrations of nitrogen dioxide (NO_2_); data for the earliest year when the participant attended the assessment centre was used. We used the World Health Organization (WHO) guideline threshold of 40 µg/m^3^ (annual mean) which was set as the level for protecting the public from the health effects of gaseous pollutants, with values ≥40 µg/m^3^ assigned as high;[30]; 8) serum 25-hydroxyvitamin D (25(OH)D) concentration, a measure of vitamin D status, was measured at a central laboratory and further details of biochemical assays are provided elsewhere (http://biobank.ctsu.ox.ac.uk/crystal/refer.cgi?id=5636). In line with the recommendation to maintain sufficient amounts of blood vitamin D levels at ≥25 nmol/L in the vast majority (97.5%) of the population in the UK,[31] participants were grouped as clinically deficient (<25 nmol/L), or sufficient (≥25 nmol/L) based on their 25(OH)D level; 9) cardiorespiratory fitness, defined by self-reported walking pace (slow walking pace, or steady-brisk walking pace [32]); and 10) the inflammatory marker high sensitivity C-reactive protein (CRP, normal [<3 mg/L] or high [≥3 mg/L]) based on clinical recommendations.[33]

### Statistical analyses

We examined the baseline descriptive characteristics of the UK Biobank cohort among participants with and without severe SARS-CoV-2 infection. The age at the time of SARS-CoV-2 test was calculated using the baseline date and the test date. Continuous variables were presented as means and standard deviations, and the categorical variables presented as counts and percentages. Complete case framework was used for all analyses. A heat map was created to illustrate the pattern of the most common clusters of multimorbidity by severe SARS-CoV-2 infection and the remaining cohort. The proportion and date of those with severe SARS-CoV-2 infection by multimorbidity was also illustrated to understand the pattern over the time of the pandemic.

The association between multimorbidity and severe SARS-CoV-2 infection was estimated using logistic regression models. The odds ratio (OR) and 95% confidence intervals (CI) were reported comparing multimorbidity versus no multimorbidity (reference). Models were adjusted for age at test, sex, ethnicity, deprivation, smoking status, BMI, air pollution, serum 25‐hydroxyvitamin D, cardiorespiratory fitness, serum C-reactive protein, season at blood draw [categorised by the months: spring (March, April, May), summer (June, July, August), autumn (September, October, November), and winter (December, January, February)], and regular intake of vitamin D supplement (yes or no). Stratified analyses were used for the ten potential effect modifiers, and we tested for interaction effects using a Likelihood ratio test and the related P-value.

In sensitivity analyses to assess the robustness of our results, we repeated the stratified analyses using 3 or more comorbid conditions. Moreover, we further stratified the analyses using four 25‐hydroxyvitamin D levels (<25, 25-50, 50-75, >75 nmol/L) and also included a more recent testing phase for 25‐hydroxyvitamin D levels; and we used the air pollution level markers NO_2_ and PM 2.5 at the last recorded measure. A P-value less than 0.05 was considered statistically significant. All analyses were performed in Stata version 16.0.

## RESULTS

### Participant characteristics

In 360,283 UK Biobank participants, 1,706 (0.5%) had severe SARS-CoV-2 infection. The median [range] age was 68 [48-85] years and most participants were White (94.5%). Compared with participants without severe SARS-CoV-2 infection, those with severe infection were older (≥ 60 years: 86% vs. 77%), mainly men (58% vs. 46%), more likely to live in deprived areas (60% vs. 50%), with higher mean BMI (28.6 kg/m^2^ vs 27.4 kg/m^2^), were current or previous smokers, had vitamin D deficiency (16% vs.12%), and more frequently reported a slow walking pace (17% vs. 7%); ***Table1***.

**Table 1.**
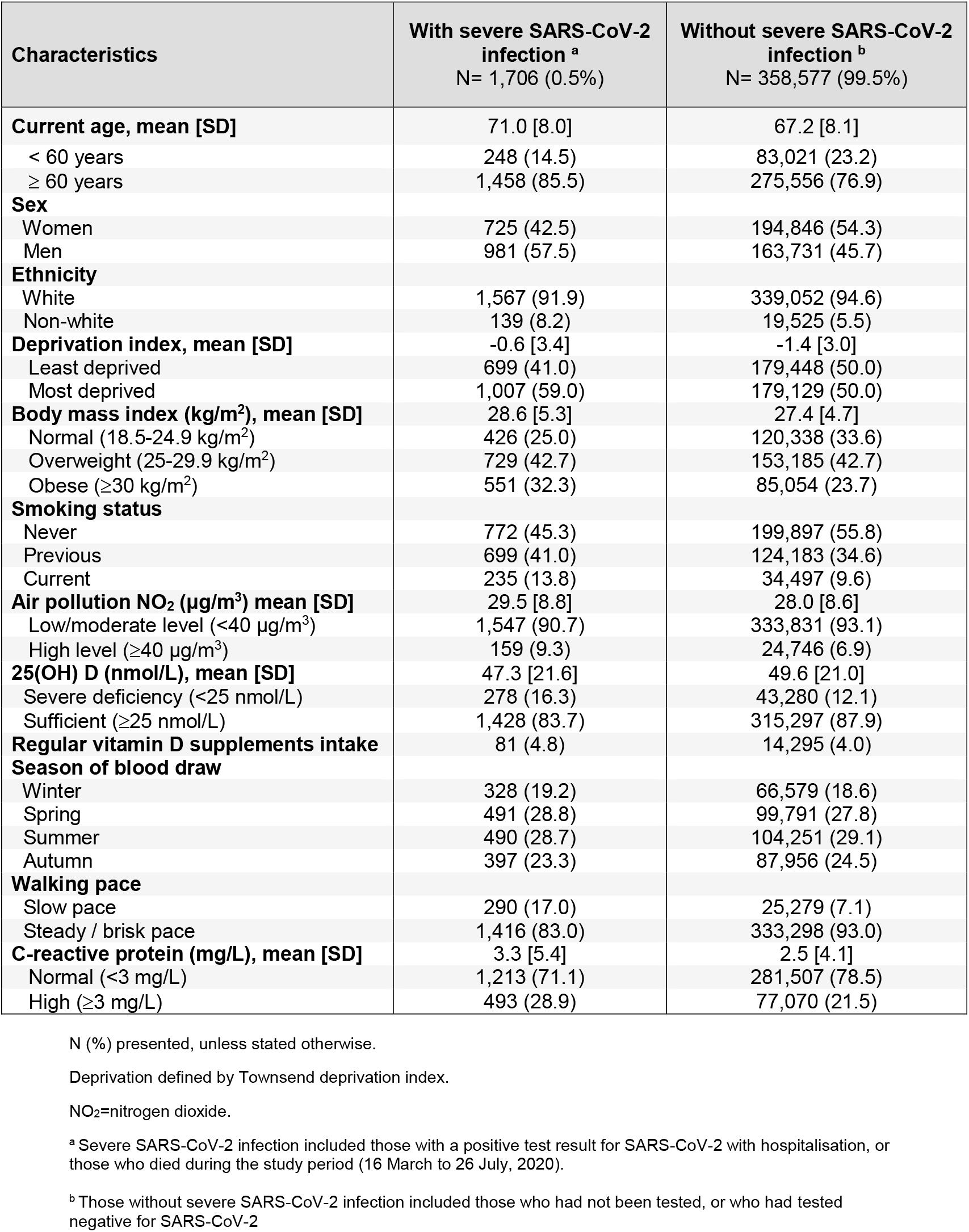
Characteristics of UK Biobank participants.

| Characteristics | With severe SARS-CoV-2 infection <sup>a</sup><br>N= 1,706 (0.5%) | Without severe SARS-CoV-2 infection <sup>b</sup><br>N= 358,577 (99.5%) |
| --- | --- | --- |
| **Current age, mean [SD]** | 71.0 [8.0] | 67.2 [8.1] |
| < 60 years | 248 (14.5) | 83,021 (23.2) |
| ≥ 60 years | 1,458 (85.5) | 275,556 (76.9) |
| **Sex** |  |  |
| Women | 725 (42.5) | 194,846 (54.3) |
| Men | 981 (57.5) | 163,731 (45.7) |
| **Ethnicity** |  |  |
| White | 1,567 (91.9) | 339,052 (94.6) |
| Non-white | 139 (8.2) | 19,525 (5.5) |
| **Deprivation index, mean [SD]** | -0.6 [3.4] | -1.4 [3.0] |
| Least deprived | 699 (41.0) | 179,448 (50.0) |
| Most deprived | 1,007 (59.0) | 179,129 (50.0) |
| **Body mass index (kg/m<sup>2</sup>), mean [SD]** | 28.6 [5.3] | 27.4 [4.7] |
| Normal (18.5-24.9 kg/m<sup>2</sup>) | 426 (25.0) | 120,338 (33.6) |
| Overweight (25-29.9 kg/m<sup>2</sup>) | 729 (42.7) | 153,185 (42.7) |
| Obese (≥30 kg/m<sup>2</sup>) | 551 (32.3) | 85,054 (23.7) |
| **Smoking status** |  |  |
| Never | 772 (45.3) | 199,897 (55.8) |
| Previous | 699 (41.0) | 124,183 (34.6) |
| Current | 235 (13.8) | 34,497 (9.6) |
| **Air pollution NO<sub>2</sub> (µg/m<sup>3</sup>) mean [SD]** | 29.5 [8.8] | 28.0 [8.6] |
| Low/moderate level (<40 µg/m<sup>3</sup>) | 1,547 (90.7) | 333,831 (93.1) |
| High level (≥40 µg/m<sup>3</sup>) | 159 (9.3) | 24,746 (6.9) |
| **25(OH) D (nmol/L), mean [SD]** | 47.3 [21.6] | 49.6 [21.0] |
| Severe deficiency (<25 nmol/L) | 278 (16.3) | 43,280 (12.1) |
| Sufficient (≥25 nmol/L) | 1,428 (83.7) | 315,297 (87.9) |
| **Regular vitamin D supplements intake** | 81 (4.8) | 14,295 (4.0) |
| **Season of blood draw** |  |  |
| Winter | 328 (19.2) | 66,579 (18.6) |
| Spring | 491 (28.8) | 99,791 (27.8) |
| Summer | 490 (28.7) | 104,251 (29.1) |
| Autumn | 397 (23.3) | 87,956 (24.5) |
| **Walking pace** |  |  |
| Slow pace | 290 (17.0) | 25,279 (7.1) |
| Steady / brisk pace | 1,416 (83.0) | 333,298 (93.0) |
| **C-reactive protein (mg/L), mean [SD]** | 3.3 [5.4] | 2.5 [4.1] |
| Normal (<3 mg/L) | 1,213 (71.1) | 281,507 (78.5) |
| High (≥3 mg/L) | 493 (28.9) | 77,070 (21.5) |

N (%) presented, unless stated otherwise.
Deprivation defined by Townsend deprivation index.
NO<sub>2</sub>=nitrogen dioxide.
<sup>a</sup> Severe SARS-CoV-2 infection included those with a positive test result for SARS-CoV-2 with hospitalisation, or those who died during the study period (16 March to 26 July, 2020).
<sup>b</sup> Those without severe SARS-CoV-2 infection included those who had not been tested, or who had tested negative for SARS-CoV-2

### Pattern of multimorbidity

The most prevalent pre-existing multimorbidity index conditions in those with severe SARS-CoV-2 infection were hypertension (39.6%), asthma (13.3%), diabetes (10.0%), cancer (9.9%), and angina (6.5%; ***Table 2***). There was a higher prevalence of hypertension (39.6% vs. 25.3%) and diabetes (10% vs. 3.9%) in individuals with severe SARS-CoV-2 infection compared to those without. The prevalence of multimorbidity (2 or more conditions) was over double in those with severe SARS-CoV-2 infection (25.3%) compared to those without severe SARS-CoV-2 infection (11.6%; ***Table 2***), where the most common clusters were found in those with both stroke and hypertension (79% of those with stroke had hypertension), diabetes and hypertension (72% of those with diabetes), CKD and hypertension (68% of those with CKD), angina and hypertension (67% of those with angina; ***Figure 1***). The trend of severe SARS-CoV-2 infection cases in this study peaked at the end of March and beginning of April 2020, and those with multimorbidity had a consistently higher proportion of cases during the period compared to those without multimorbidity; ***Figure 2***.

**Table 2.** Multimorbidity index conditions among participants in the UK Biobank Study.

| Multimorbidity index conditions | With severe SARS-CoV-2 infection <sup>a</sup><br>N= 1,706 (0.5%) | Without severe SARS-CoV-2 infection <sup>b</sup><br>N= 358,577 (99.5%) |
| --- | --- | --- |
| Hypertension | 676 (39.6) | 90,747 (25.3) |
| Asthma | 226 (13.3) | 41,382 (11.5) |
| Diabetes | 170 (10.0) | 14,079 (3.9) |
| Cancer | 179 (10.5) | 27,108 (7.6) |
| Angina | 111 (6.5) | 10,037 (2.8) |
| Myocardial infarction | 102 (6.0) | 7,096 (2.0) |
| Chronic kidney disease | 93 (5.5) | 7,188 (2.0) |
| Stroke | 47 (2.8) | 3,957 (1.1) |
| Atrial fibrillation | 26 (1.5) | 2,508 (0.7) |
| Chronic obstructive pulmonary disease | 14 (0.8) | 929 (0.3) |
| Heart failure | 5 (0.3) | 146 (0.04) |
| Peripheral vascular disease | 4 (0.2) | 595 (0.2) |
| **Number of multimorbidity index conditions** |  |  |
| None | 683 (40.0) | 206,012 (57.5) |
| One | 592 (34.7) | 111,153 (31.0) |
| Two | 293 (17.2) | 32,059 (8.9) |
| Three | 96 (5.6) | 7,341 (2.1) |
| Four | 27 (1.6) | 1,646 (0.5) |
| Five | 12 (0.7) | 311 (0.1) |
| Six | 2 (0.1) | 47 (0.01) |
| Seven | 1 (0.1) | 7 (0.0) |
| Eight | 0 (0.0) | 1 (0.0) |

<sup>a</sup> Severe SARS-CoV-2 infection included those with a positive test result for SARS-CoV-2 with hospitalisation, or those who died during the study period (16 March to 26 July, 2020).
<sup>b</sup> Those without severe SARS-CoV-2 infection included those who had not been tested, or who had tested negative for SARS-CoV-2.

**Figure 1.**
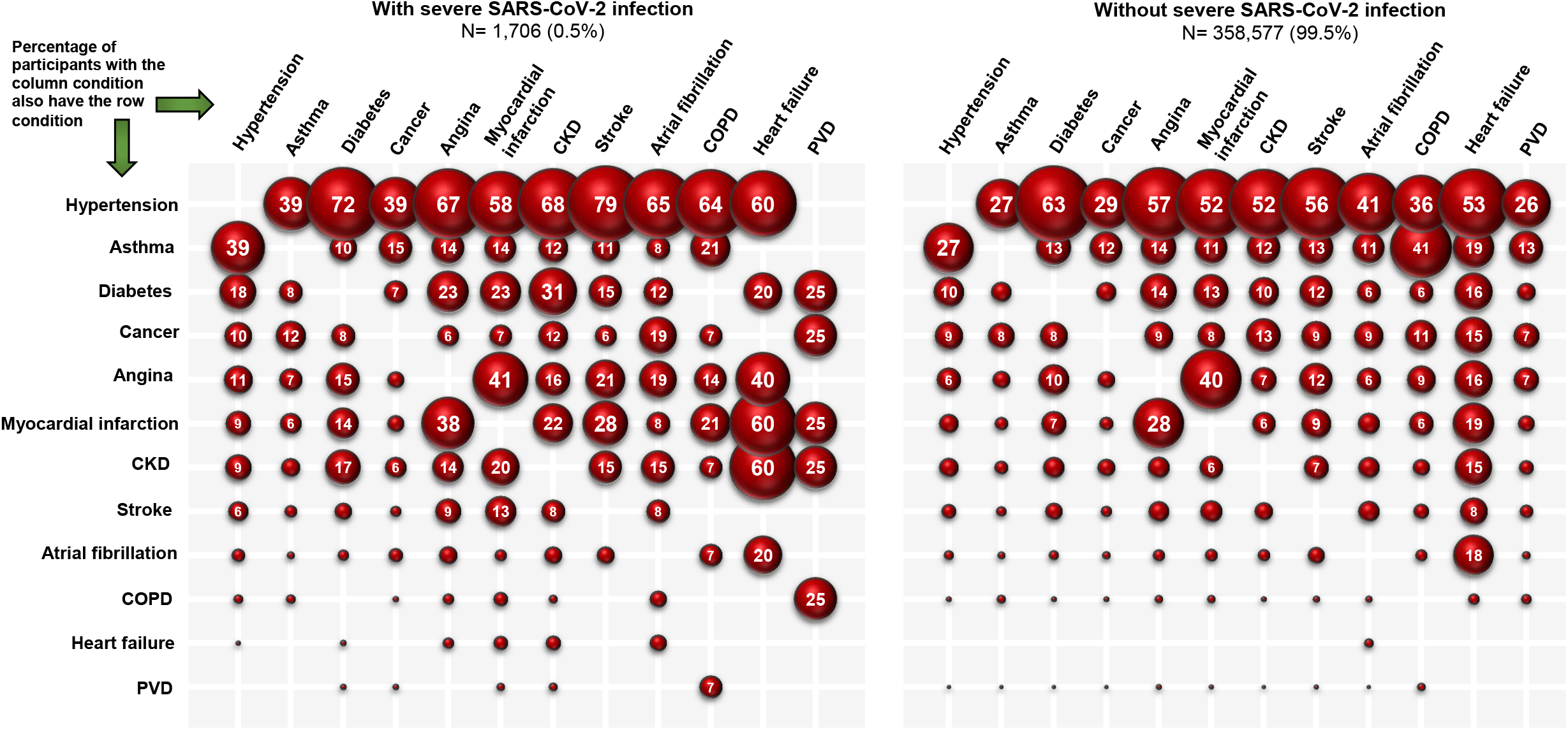
Two most common pre-existing multimorbidity index conditions in UK Biobank participants with and without severe SARS-CoV-2 infection (March 16 to July 26, 2020) CKD=chronic kidney disease; CPD=chronic obstructive pulmonary disease; PVD= peripheral vascular disease

**Figure 2.**
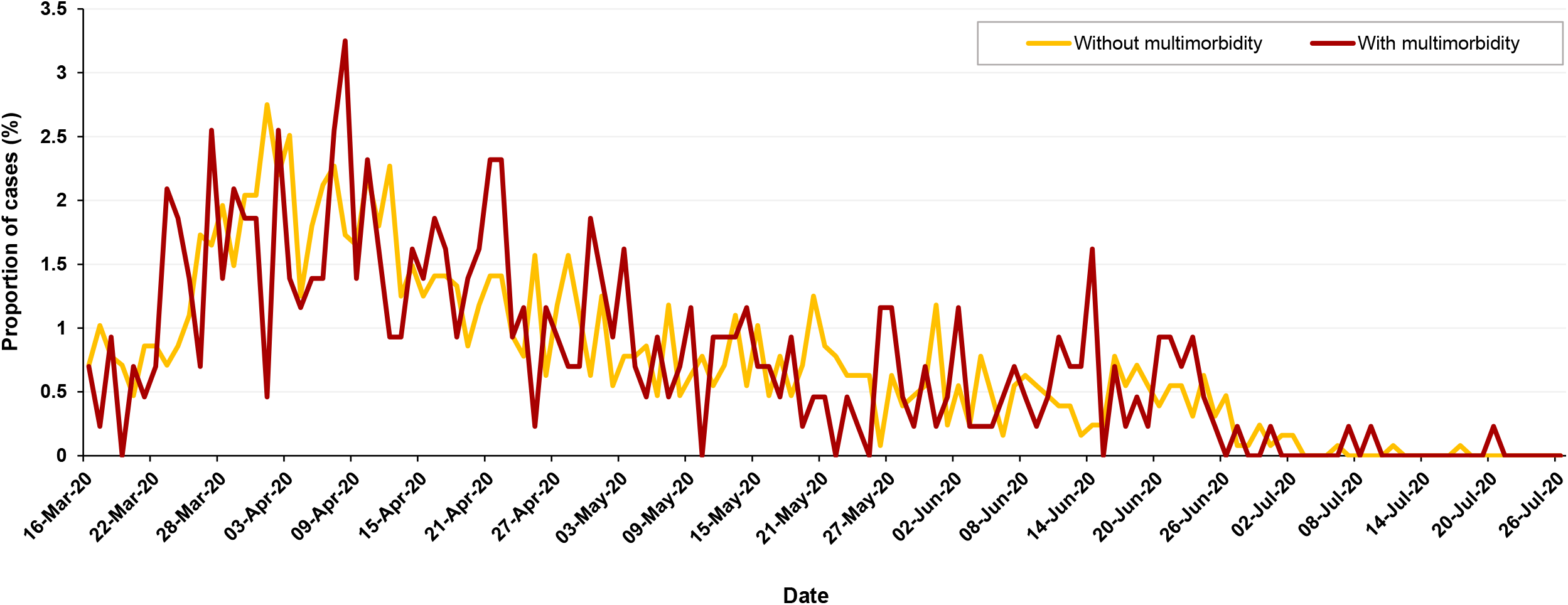
Comparing the proportion and date of those diagnosed with severe SARS-CoV-2 infection (hospitalisation or death, N=1,706) by pre-existing multimorbidity index conditions (≥ 2 conditions) in the UK Biobank. Without multimorbidity (n=1, 275); with multimorbidity (n=431). Multimorbidity index conditions include: angina, asthma, atrial fibrillation, cancer, chronic kidney disease, chronic obstructive pulmonary disease, diabetes mellitus, heart failure, hypertension, myocardial infarction, peripheral vascular disease, and stroke.

### Risk of severe SARS-CoV-2 infection

The risk of severe SARS-CoV-2 infection increased with the number of pre-existing multimorbidity index conditions, illustrated by the higher odds ratios across the number of increasing conditions (***Table 3)***. The overall combined effect in those with multimorbidity (2 or more conditions) was associated with around 2.6 times greater risk of severe SARS-CoV-2 infection compared to those without multimorbidity (crude OR: 2.56 [2.32, 2.89]); and, in the adjusted model, nearly 2 times greater (adjusted OR: 1.91 [1.70, 2.15]), ***Table 3***. The odds ratios remained generally consistent with the overall associations when estimated across strata of effect modifiers (***Figure 3***). The association was slightly stronger in men compared to women. It was also apparent that the risk differed across the most common clusters of multimorbidity (***Table 4***) with clusters containing asthma & hypertension having the lowest risk (adjusted OR 1.29 [1.03, 1.60]) and those containing CKD & diabetes having the highest risk (adjusted OR 4.93 [3.36, 7.22]).

**Table 3.** Association between the number of multimorbidity index conditions and the risk of severe SARS-CoV-2 infection (N=360,283): the UK Biobank Study

| **Risk of severe SARS-CoV-2 infection** (hospitalisation or death) | **Crude OR (95% CI)** | **Adjusted OR (95% CI)** |
| --- | --- | --- |
| ≥ 1 vs. 0 (reference) | 2.02 (1.84, 2.23) | 1.65 (1.49, 1.82) |
| ≥ 2 vs. <2 (reference) | 2.59 (2.32, 2.89) | 1.91 (1.70, 2.15) |
| ≥ 3 vs. <3 (reference) | 3.29 (2.76, 3.92) | 2.02 (1.68, 2.43) |
| ≥ 4 vs. <4 (reference) | 4.47 (3.28, 6.09) | 2.25 (1.64, 3.09) |

**Table 4.** Association between the most common co-occurring multimorbidity index conditions and the risk of severe SARS-CoV-2 infection: UK Biobank (N=360,283)

| **Risk of severe SARS-CoV-2 infection**<br>(hospitalisation or death) | **Adjusted OR (95% CI)** |
| --- | --- |
| Asthma & hypertension (n=11,242) | 1.29 (1.03, 1.60) |
| Diabetes & hypertension (n=9,025) | 1.92 (1.59, 2.33) |
| Cancer & hypertension (n=7,783) | 1.58 (1.24, 2.01) |
| Angina & hypertension (n=5,802) | 1.66 (1.31, 2.11) |
| Myocardial infarction & hypertension (n=3,763) | 1.98 (1.51, 2.59) |
| CKD & hypertension (n=3,817) | 2.58 (2.00, 3.34) |
| Stroke & hypertension (n=2,236) | 2.16 (1.55, 3.02) |
| Atrial fibrillation & hypertension (n=1,052) | 2.44 (1.50, 3.96) |
| COPD & hypertension (n=343) | 2.70 (1.38, 5.28) |
| Heart failure & hypertension (n=81) | 4.47 (1.40, 14.29) |
| CKD & diabetes (n=713) | 4.93 (3.36, 7.22) |
| Myocardial infarction & angina (n=2,814) | 1.78 (1.30, 2.43) |

Reference is those without the stated two co-occurring conditions (i.e. with asthma and hypertension vs. without asthma and hypertension [reference]).
OR=odds ratio; CI=confidence interval; CKD=chronic kidney disease; CPD=chronic obstructive pulmonary disease.
Most common co-occurring multimorbidity conditions illustrated in Figure 1.

**Figure 3.**
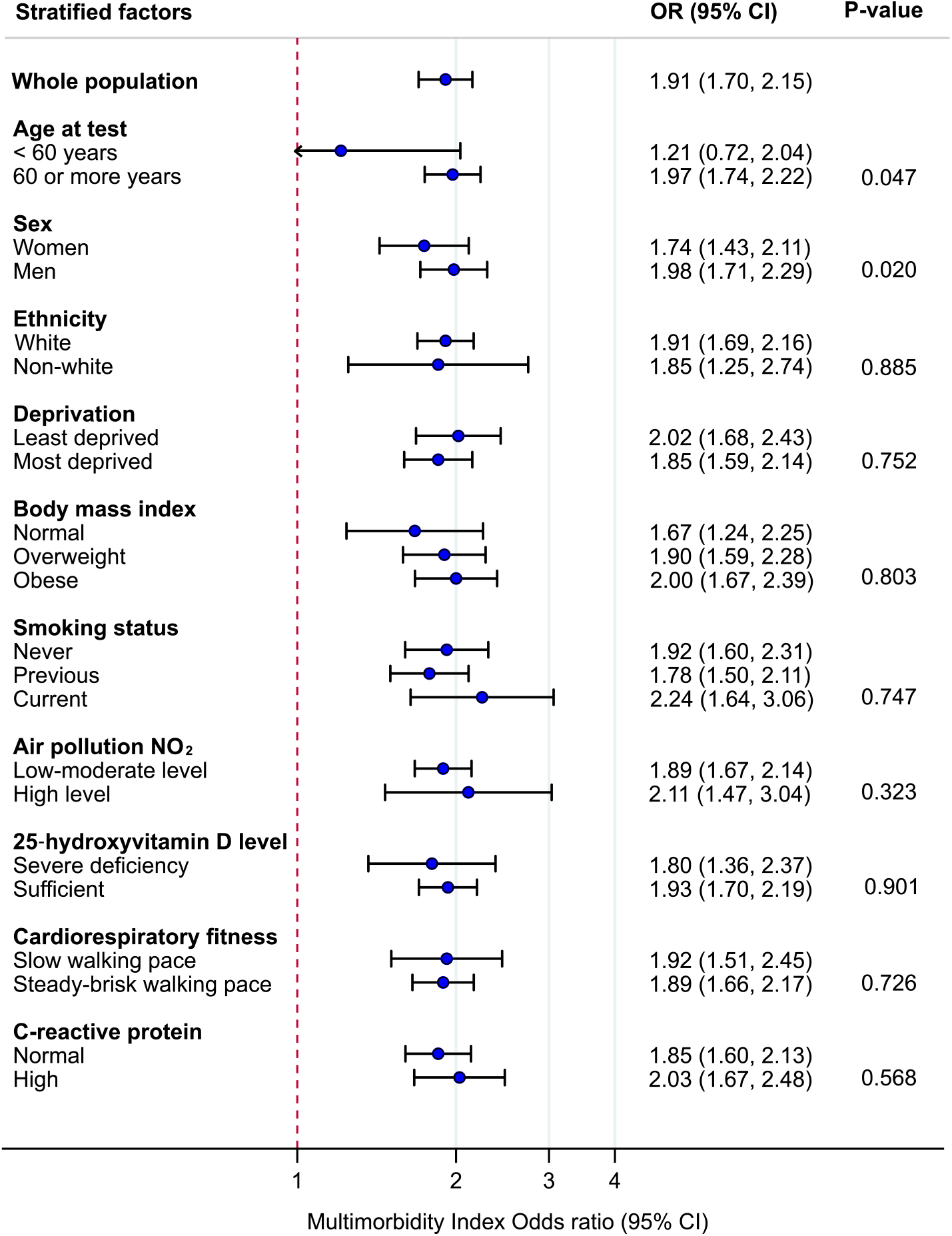
Association between the multimorbidity index and risk of severe SARS-CoV-2 infection across strata of effect modifiers (N=360,283) in the UK Biobank Study. Reference is the group without multimorbidity (<2 conditions). OR=odds ratio; CI=confidence interval; NO_2_=nitrogen dioxide. Multimorbidity index conditions include: angina, asthma, atrial fibrillation, cancer, chronic kidney disease, chronic obstructive pulmonary disease, diabetes mellitus, heart failure, hypertension, myocardial infarction, peripheral vascular disease, and stroke. Models adjusted for age at test, sex, ethnicity, deprivation, smoking status, body mass index, air pollution, 25‐hydroxyvitamin D, cardiorespiratory fitness, C-reactive protein, season at blood draw, and regular intake of Vitamin D supplement.

In sensitivity analyses, the results were similar when examining the association between the multimorbidity index using 3 or more comorbid conditions and the risk of severe SARS-CoV-2 infection (***Additional file: Table S2***), however the interaction effect between men and women was non-significant. The results remained consistent over time, when assessing the 25OH-vitamin D levels at a later phase and when using the last recorded air pollution NO_2_ and PM 2.5 levels (***Additional file: Table S3***).

## DISCUSSION

Our review of the published literature indicated that the most commonly reported chronic conditions for severe SARS-CoV-2 infection were cardiometabolic diseases. Data from the UK Biobank study using the multimorbidity index conditions showed that 25% of those with severe SARS-CoV-2 infection had multimorbidity (2 or more conditions). The major clusters were stroke and hypertension; diabetes and hypertension; CKD and hypertension; and angina and hypertension. Over the study period the risk of severe SARS-CoV-2 infection was consistently higher for those with multimorbidity compared to those without multimorbidity. Moreover, our results highlight that the greater risk of severe infection was consistent with that of the overall effect of multimorbidity when stratifying by the potential effect modifiers. The only exception was when stratifying by sex, with a greater magnitude of association between multimorbidity and severe infection in men compared with women.

To our knowledge, this is the first study to develop a multimorbidity index using the most common conditions from the current COVID-19 literature. There have been three previously published studies that assessed the association between multimorbidity and the risk of SARS-CoV-2 infection. The first was an online questionnaire study by 26 hospitals across Italy undertaken in March 9 to April 9, 2020, with a total of 1,591 participants admitted with SARS-CoV-2 infection (188 non-survivors, 1,403 survivors).[14] The study used the Charles Comorbidity Index (CCI) which takes into account age and 19 conditions in an algorithm to predict 10-year mortality. The authors found an exponential increase in the odds by each point of the CCI score.[14] Our multimorbidity index found similar results since the risk of severe SARS-CoV-2 infection increased with the number of pre-existing multimorbidity index conditions. Two studies were based in UK Biobank, however the study design and outcomes differed which resulted in inconsistent findings. One study in 4,510 UK Biobank participants between 16 March to 1 June, 2020 (1,326 tested positive for COVID-19 and 3,184 tested negative) found in adjusted analyses that the number of comorbidity groupings (0-1, 2, 3, ≥ 4) was not associated with the risk of testing positive for COVID-19 when compared to those who tested negative: the OR was 1.0 [95% CI 0.8, 1.2] comparing those with 2 conditions vs.0-1 conditions; or only slightly increased when compared to the remaining population (OR 1.3 [1.1, 1.5]).[13] However, this study used 43 different conditions to define multimorbidity, and compared only those testing positive compared to negative. The other UK Biobank study, between 16 March and 18 May, 2020 (1,324 tested positive for COVID-19 and 426,875 remaining cohort), used Poisson regression analyses reporting that those with multimorbidity (defined using 43 conditions) had a 48% higher risk for a positive test for COVID-19 (RR 1.48 [95% CI 1.28, 1.71], P<0.01). Although this study also reported that those with cardiometabolic multimorbidity, compared with those without, had a 77% higher risk for a positive test (RR 1.77 [1.46, 2.15] P<0.01),[15] the outcome was based on testing positive in any setting (hospital or outpatient, pillar 1 and 2), which may introduce testing bias within UK Biobank. A recent article illustrated how the same dataset can produce apparently paradoxical findings for the same outcome.[34] For this reason, our study examined harder outcomes for those with severe SARS-CoV-2 infection; hospitalisation or death during the pandemic compared to the remaining cohort. Moreover, the three studies to date have only presented the number of conditions but have not examined the patterns of multimorbidity, i.e. the type of the two most common co-occurring conditions found in those with severe SARS-CoV-2 infection, nor the rate of severe infection or mortality in people with multimorbidity over the period of the pandemic.

Hypertension was found to be the most prevalent condition (40%) in those with severe SARS-CoV-2 infection, as in a number of previously published studies. Yet, when examined by clusters, we found that hypertension mainly co-existed with other conditions (i.e. diabetes, stroke, CKD). Thus, certain clusters of the multimorbidity index are more frequent than other clusters. For this reason, we believe that the classification of those with pre-existing multimorbidity index conditions is crucial to provide prognostic information to tailor effective treatment and prevent severe outcomes in common chronic conditions. Further research is required to understand the biological mechanisms involved between these multimorbidity clusters and the increased risk of severe SARS-CoV-2 infection, particularly the effects on the immune system.

Multimorbidity has been associated with premature mortality, lower quality of life, polypharmacy, and fragmented care.[10] However, the current pandemic has placed additional risks on individuals with multimorbidity. A global survey evaluating the impact of COVID-19 on routine care for chronic diseases during 31 March to 23 April 2020 found that most healthcare professionals (67%) identified moderate or severe effects on their patients due to changes in healthcare services since the outbreak; and 80% reported the mental health of their patients worsened during COVID-19.[35] Moreover, many low and middle-income countries have overstretched healthcare systems and the pandemic has further overwhelmed them.

A key strength of this study is the overall large sample size which allowed us to investigate whether the associations were heterogeneous across clinically relevant effect modifiers. We employed a comprehensive list of effect modifiers, extending past research and enabling a greater understanding of the association between multimorbidity and severe SARS-CoV-2 infection. Moreover, we performed a literature review to derive an evidence based COVID-19 relevant multimorbidity index which can be applied to other populations.[5-8, 20-29] This study has some limitations. Issues of the low response rate (∼5%) and selection bias, such as slightly higher representation of participants from affluent groups may suggest that the UK Biobank sample is not well representative of the UK population, and have been discussed previously.[36] However, participants may not need to be representative of the target populations when estimating relative risks, as empirically demonstrated for UK Biobank.[37] The characteristics we examined were recorded at recruitment, representing data collected in the past at study baseline, but we conducted additional sensitivity analyses using the follow-up data for 25‐hydroxyvitamin D levels and the last recorded air pollution (NO_2_) data, showing consistent results over time. The data on multimorbidity were appraised only at baseline but there may have been a change in the number of multimorbidity index conditions since recruitment which were not accounted for and may lead to under or overestimation of effect. We were unable to assess the severity of disease conditions but our new multimorbidity index definition included conditions that were most relevant to COVID-19. Another limitation is the lack of detail of clinical severity of those with severe SARS-CoV-2 infection: while we have accounted for hospitalisation or death during the pandemic, further details of adverse COVID-19 outcomes such as length of hospital stay, intensive care admission, or mechanical ventilation, could provide further information. Ten potential effect modifiers were selected,[4] although there are other factors which could possibly modify the association between multimorbidity and risk of severe SARS-CoV-2 infection but were unaccounted for, such as household size, occupation, and understanding and behaviour for social distancing, shielding, wearing facemasks and hand hygiene. Finally, this was an observational study, which limits our ability to establish causality.

In conclusion, our results indicate that multimorbidity is associated with a significantly higher risk of severe SARS-CoV-2 infection across multiple patient characteristics. Stratifying the risk according to the presence of multimorbidity, and reinforcing initiatives and resources to reduce the risk in this vulnerable group of individuals should be considered as priorities in the current coronavirus pandemic.

## Data Availability

The data that support the findings of this study are available from UK Biobank project site, subject to registration and application process. Further details can be found at https://www.ukbiobank.ac.uk. Data on patients with COVID-19 is from an updated database which has not been reported in other submissions.
The UK Biobank protocol https://www.ukbiobank.ac.uk/wp-content/uploads/2011/11/UK-Biobank-Protocol.pdf
The statistical codes are available at GitHub yc244.
https://github.com/yc244/Patterns-of-Multimorbidity-and-Risk-of-Severe-SARS-CoV-2-Infection-

https://www.ukbiobank.ac.uk

https://www.ukbiobank.ac.uk/wp-content/uploads/2011/11/UK-Biobank-Protocol.pdf

https://github.com/yc244/Patterns-of-Multimorbidity-and-Risk-of-Severe-SARS-CoV-2-Infection-

## List of Abbreviations

BMI: Body mass index
CI: Confidence intervals
CKD: Chronic kidney disease
COPD: Chronic obstructive pulmonary disease
CRP: C-reactive protein
eGFR: Estimated Glomerular Filtration Rate
IQR: Interquartile range
NHS: National Health Service
OR: Odds ratio
RR: Relative risk
UK: United Kingdom
WHO: World Health Organization
25(OH)D: 25-hydroxyvitamin D

## Author contributions

Concept and Design: NG Forouhi, K Khunti

Statistical analysis: YV Chudasama

Statistical support: NG Forouhi, F Zaccardi, CL Gillies

Acquisition, analysis, or interpretation of data: All authors

Drafting of the manuscript: YV Chudasama

Critical revision of the manuscript for important intellectual content: All authors

## Acknowledgements

This research has been conducted using the UK Biobank Resource (Reference 36371). We acknowledge the support from the National Institute for Health Research (NIHR) Applied Research Collaboration East Midlands (ARC EM), and the NIHR Leicester Biomedical Research Centre. NGF acknowledges funding from the MRC Epidemiology Unit core support (MC_UU_12015/5), and NIHR Biomedical Research Centre Cambridge: Nutrition, Diet, and Lifestyle Research Theme (IS-BRC-1215-20014). The views expressed are those of the author(s) and not necessarily those of the NIHR or the Department of Health and Social Care.

## Ethical approval and informed consent

All participants gave written informed consent prior data collection. UK Biobank has full ethical approval from the NHS National Research Ethics Service (16/NW/0274).

## Data access and responsibility

The data that support the findings of this study are available from UK Biobank project site, subject to registration and application process. Further details can be found at https://www.ukbiobank.ac.uk.

