## Additional File for "Patterns of Multimorbidity and Risk of Severe SARS-CoV-2 Infection: an observational study in the U.K"

### Additional File: Supplementary

**Figure S1:** Flow chart of participants included in the UK Biobank Study

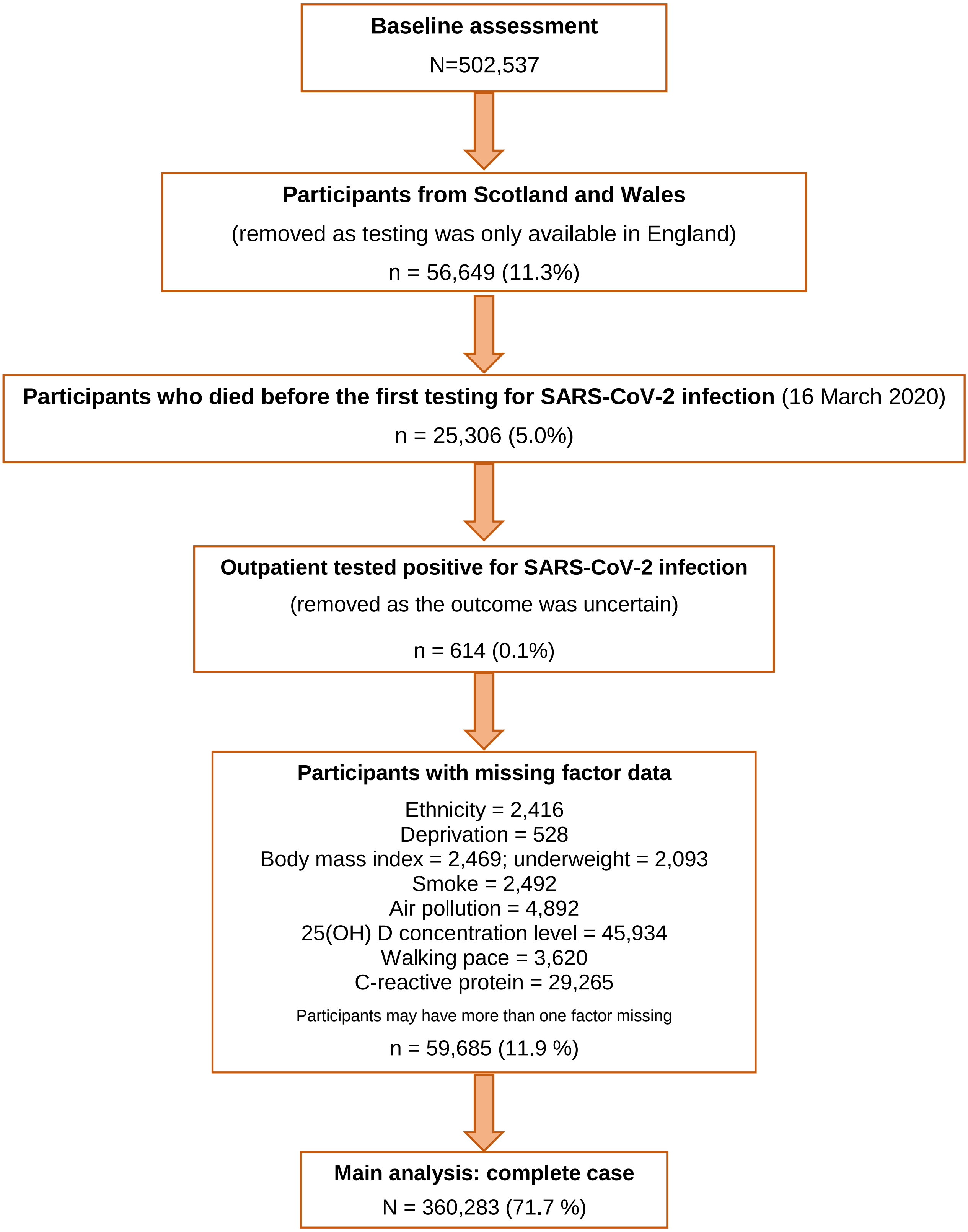

**Table S1:** Literature search on the most common pre-existing comorbidities in patients with severe SARS-CoV-2 infection

| Reference | Type of study and date | Study population | Most common pre-existing comorbidities associated with patients with severe SARS-CoV-2 infection (hospitalised) |
|---|---|---|---|
| **Arentz et al, 2020 [1]** | Case series<br>February 20 to March 5, 2020 | 21 critically ill patients with COVID-19 in Washington State, United States | 1. Chronic kidney disease<br>2. Heart failure<br>3. Diabetes<br>4. Chronic obstructive pulmonary disease<br>5. Obstructive sleep apnoea<br>6. Asthma |
| **Du et al. 2020 [2]** | Prospective cohort study,<br>25 December 2019, to 7 February 2020 | 179 patients who were hospitalised with COVID-19 to Wuhan Pulmonary Hospital, China | 1. Hypertension<br>2. Diabetes mellitus<br>3. Cardiovascular or cerebrovascular diseases<br>4. Chronic digestive disorders<br>5. Tuberculosis<br>6. Cancer, malignancy<br>7. Peripheral vascular disease |
| **Emami et al. 2020 [3]** | Systematic review and meta-analysis<br>Until 15 February 2020<br>10 articles | 3,403 hospitalised patients with COVID-19 | 1. Hypertension<br>2. Cardiovascular diseases<br>3. Diabetes mellitus<br>4. Chronic obstructive pulmonary disease<br>5. Cancer, malignancy<br>6. Chronic kidney disease |
| **Grasselli et al, 2020 [4]** | Retrospective case series<br>February 20 to March 18 2020 | 1591 critically ill patients admitted to ICUs in Lombardy, Italy | 1. Hypertension<br>2. Cardiovascular disease<br>3. Hypercholesterolemia<br>4. Diabetes, type 2<br>5. Cancer, malignancy<br>6. Chronic obstructive pulmonary disease<br>7. Chronic liver disease<br>8. Chronic kidney disease |
| **Guan et al. 2020 [5]** | Retrospective case study,<br>11 December 2019, to 31 January 2020 | 1590 laboratory confirmed hospitalised patients from 575 hospitals in 31 provinces/autonomous regions/provincial municipalities across mainland China | 1. Hypertension<br>2. Cardiovascular or cerebrovascular diseases<br>3. Diabetes mellitus<br>4. Hepatitis B infection<br>5. Chronic kidney disease<br>6. Cancer, malignancy |

| Reference | Type of study and date | Study population | Most common pre-existing comorbidities associated with patients with severe SARS-CoV-2 infection (hospitalised) |
|---|---|---|---|
| **Ji et al, 2020 [6]** | Nationwide retrospective case-control study<br>Until May 15 2020 | Severe cases were 954 of 7,341, Korea | 1. Hypertension<br>2. Diabetes mellitus<br>3. Chronic lower respiratory disease<br>4. Chronic renal failure |
| **Li X et al, 2020 [7]** | Retrospective study<br>January 26 to February 5 2020 | 548 patients as severe cases on admission, Tongji Hospital, Wuhan, China | 1. Hypertension<br>2. Diabetes<br>3. Asthma<br>4. Coronary heart disease<br>5. Tuberculosis<br>6. Chronic obstructive pulmonary disease<br>7. Cancer, tumour<br>8. Chronic kidney disease<br>9. Hepatitis B |
| **Myers et al. 2020 [8]** | Retrospective cohort study,<br>March 1 2020, to March 31 2020 | 377 were treated as inpatients and 113 were treated in the ICU, in 21 hospitals, California, United States | 1. Hypertension<br>2. Diabetes mellitus<br>3. Chronic kidney disease<br>4. Chronic obstructive pulmonary disease or asthma<br>5. Heart failure<br>6. Liver cirrhosis<br>7. Cancer, malignancy |
| **Petrilli et al, 2020 [9]** | Prospective cohort study<br>1 March 2020 and 8 April 2020 | 2741 were admitted to hospital, New York City and Long Island, United States | 1. Hypertension<br>2. Cardiovascular disease<br>3. Asthma or chronic obstructive pulmonary disease<br>4. Diabetes<br>5. Cancer<br>6. Chronic kidney disease |
| **Q et al. 2020 [10]** | Retrospective cohort study,<br>January 30 2020, to February 11 2020 | 108 adult patients with COVID-19 were hospitalised in the Dabieshan Medical Center, Huanggang, China | 1. Hypertension<br>2. Diabetes mellitus<br>3. Chronic obstructive pulmonary disease<br>4. Cardiovascular disease<br>5. Chronic liver disease<br>6. Cancer |

| Reference | Type of study and date | Study population | Most common pre-existing comorbidities associated with patients with severe SARS-CoV-2 infection (hospitalised) |
|---|---|---|---|
| **Richardson et al. 2020 [11]** | Case series,<br>March 1 2020, to April 4 2020 | 5,700 hospitalised patients with COVID-19, in 12 hospitals across New York, United States | 1. Hypertension<br>2. Cardiovascular disease<br>3. Obesity<br>4. Diabetes mellitus<br>5. Cancer |
| **Yang J et al, 2020 [12]** | Systematic review and meta-analysis<br>Until 25 February 2020<br>7 articles | 1,576 infected patients from hospitals in China | 1. Hypertension<br>2. Diabetes mellitus<br>3. Respiratory system disease<br>4. Cardiovascular disease |
| **Yang X et al. 2020 [13]** | Retrospective study<br>Before 31 January 2020 | 52 critically ill adult patients with SARS-CoV-2 pneumonia who were admitted to the intensive care unit of Wuhan Jin Yin-tan hospital,<br>China | 1. Cerebrovascular disease<br>2. Diabetes mellitus<br>3. Chronic cardiac disease<br>4. Chronic pulmonary disease |
| **Zhou et al. 2020 [14]** | Retrospective, multicentre cohort study<br>Before 31 January 2020 | 191 patients with COVID-19 (135 from Jinyintan Hospital and 56 from Wuhan Pulmonary Hospital), China | 1. Hypertension<br>2. Diabetes mellitus<br>3. Coronary heart disease<br>4. Chronic obstructive lung disease<br>5. Cancer, Carcinoma<br>6. Chronic kidney disease |

Google Scholar and PubMed searches for studies in published in English were carried out with the terms “comorbidity”; “severe SARS-CoV-2”; or “COVID-19 hospitalisation” on 3$^{rd}$ July 2020. In the table we reported the studies we deemed most relevant. We did not include studies that were already considered in the systematic reviews.

**Table S2.** Association between multimorbidity index using 3 or more conditions and risk of severe SARS-CoV-2 infection: the UK Biobank Study

| **Risk of severe SARS-CoV-2 infection** (hospitalisation or death) | **OR (95% CI)** | **P-value** |
|---|---|---|
| **Age at test** | | |
| **< 60 years** (n=83,269) | 2.01 (0.73, 5.52) | 0.790 |
| **≥ 60 years** (n=277,014) | 2.00 (1.66, 2.42) | |
| **Sex** | | |
| **Women** (n=195,571) | 1.81 (1.28, 2.57) | 0.106 |
| **Men** (n=164,712) | 2.03 (1.63, 2.53) | |
| **Ethnicity** | | |
| **White** (n=340,619) | 1.94 (1.60, 2.36) | 0.330 |
| **Non-white** (n=19,664) | 2.75 (1.60, 4.72) | |
| **Deprivation** | | |
| **Least deprived** (n=180,147) | 2.65 (1.98, 3.56) | 0.070 |
| **Most deprived** (n=180,136) | 1.72 (1.36, 2.18) | |
| **Body mass index** | | |
| **Normal** (n=120,764) | 1.13 (0.55, 2.30) | 0.340 |
| **Overweight** (n=153,914) | 2.25 (1.67, 3.04) | |
| **Obese** (n=85,605) | 2.05 (1.59, 2.62) | |
| **Smoke** | | |
| **Never** (n=200,669) | 2.06 (1.49, 2.83) | 0.577 |
| **Previous** (n=124,882) | 2.00 (1.56, 2.56) | |
| **Current** (n=34,732) | 1.71 (1.00, 2.92) | |
| **Air pollution ($NO_2$)** | | |
| **Low/moderate level** (n=335,378) | 2.00 (1.64, 2.43) | 0.467 |
| **High level** (n=24,905) | 2.12 (1.25, 3.60) | |
| **25-hydroxyvitamin D levels** | | |
| **Severe deficiency** (n=43,558) | 1.22 (0.76, 1.95) | 0.036 |
| **Sufficient** (n=316,725) | 2.24 (1.84, 2.74) | |
| **Cardiorespiratory fitness** | | |
| **Slow walking pace** (n=25,569) | 2.00 (1.51, 2.64) | 0.772 |
| **Steady-brisk walking pace** (n=334,714) | 1.98 (1.55, 2.54) | |
| **C-reactive protein level** | | |
| **Normal** (n=282,720) | 2.24 (1.78 2.81) | 0.121 |
| **High** (n=77,563) | 1.68 (1.23, 2.30) | |

Reference is the group without multimorbidity.
OR=odds ratio; CI=confidence interval; $NO_2$=nitrogen dioxide.

Models adjusted for age at test, sex, ethnicity, deprivation, smoking status, body mass index, air pollution, 25-hydroxyvitamin D, cardiorespiratory fitness, C-reactive protein, season at blood draw, and regular intake of vitamin D supplement.

**Table S3.** Sensitivity analyses using vitamin D levels at follow-up and last recorded air pollution levels: the UK Biobank Study

| **Risk of severe SARS-CoV-2 infection** ( hospitalisation or death) | **OR (95% CI)** | |
|---|---|---|
| | **2 or more pre-existing multimorbidity index conditions** | **3 or more pre-existing multimorbidity index conditions** |
| **25-hydroxyvitamin D levels categories** | | |
| **<25 nmol/L** (n=43,558) | 1.80 (1.36, 2.37) | 1.22 (0.76, 1.94) |
| **25-50 nmol/L** (n=148,624) | 2.01 (1.69, 2.40) | 2.09 (1.59, 2.75) |
| **50-75 nmol/L** (n=125,430) | 1.84 (1.47, 2.30) | 2.71 (1.94, 3.80) |
| **≥75 nmol/L** (n=42,671) | 1.81 (1.27, 2.58) | 1.76 (0.96, 3.23) |
| **25-hydroxyvitamin D levels at follow-up *** | | |
| **Severe deficiency** (n=2,165) | 2.30 (0.75, 7.11) | 1.21 (0.14, 10.36) |
| **Sufficient** (n=12,376) | 3.14 (1.67, 5.93) | 2.92 (0.99, 8.56) |
| **Air pollution ($NO_2$) last recorded** | | |
| **Low-moderate level** (n=344,059) | 1.93 (1.71, 2.17) | 2.08 (1.72, 2.52) |
| **High level** (n=16,224) | 1.66 (1.03, 2.68) | 1.25 (0.58, 2.69) |
| **Air pollution (PM 2.5) last recorded** | | |
| **Low-moderate level** (n=192,392) | 2.08 (1.75, 2.46) | 2.50 (1.92, 3.25) |
| **High level** (n=167,891) | 1.78 (1.51, 2.08) | 1.68 (1.30, 2.18) |

Reference is the group without multimorbidity.
OR=odds ratio; CI=confidence interval; $NO_2$=nitrogen dioxide.

Models adjusted for age at test, sex, ethnicity, deprivation, smoking status, body mass index, air pollution, 25-hydroxyvitamin D, cardiorespiratory fitness, C-reactive protein, season at blood draw, and regular intake of Vitamin D supplement.

***** Model adjusted for regular intake of Vitamin D supplement at follow-up, the season of blood draw was not known at follow-up.

**Checklist S1.** Strengthening the Reporting of Observational Studies in Epidemiology (STROBE)

| | Item No | Recommendation | |
|---|---|---|---|
| **Title and abstract** | 1 | (*a*) Indicate the study's design with a commonly used term in the title or the abstract | Title and abstract |
| | | (*b*) Provide in the abstract an informative and balanced summary of what was done and what was found | Abstract, methods and findings |
| **Introduction** | | | |
| Background/rationale | 2 | Explain the scientific background and rationale for the investigation being reported | Introduction paragraph 1-3 |
| Objectives | 3 | State specific objectives, including any prespecified hypotheses | Introduction paragraph 4 |
| **Methods** | | | |
| Study design | 4 | Present key elements of study design early in the paper | Methods, Study Population |
| Setting | 5 | Describe the setting, locations, and relevant dates, including periods of recruitment, exposure, follow-up, and data collection | Methods, Study Population |
| Participants | 6 | (*a*) *Cohort study*—Give the eligibility criteria, and the sources and methods of selection of participants. Describe methods of follow-up | Methods, Study Population |
| | | (*b*) *Cohort study*—For matched studies, give matching criteria and number of exposed and unexposed | NA |
| Variables | 7 | Clearly define all outcomes, exposures, predictors, potential confounders, and effect modifiers. Give diagnostic criteria, if applicable | Methods, Multimorbidity index, Outcome measures, Effect modifiers |
| Data sources/ measurement | 8* | For each variable of interest, give sources of data and details of methods of assessment (measurement). Describe comparability of assessment methods if there is more than one group | Methods, Multimorbidity index, Outcome measures, Effect modifiers |
| Bias | 9 | Describe any efforts to address potential sources of bias | Methods, Study Population, Statistical Analysis paragraph 3 |
| Study size | 10 | Explain how the study size was arrived at | Supporting Information, Figure S1 |
| Quantitative variables | 11 | Explain how quantitative variables were handled in the analyses. If applicable, describe which groupings were chosen and why | Methods, Multimorbidity index, Effect modifiers, Statistical analysis |
| Statistical methods | 12 | (*a*) Describe all statistical methods, including those used to control for confounding | Methods, Statistical analysis |
| | | (*b*) Describe any methods used to examine subgroups and interactions | NA |
| | | (*c*) Explain how missing data were addressed | Methods, Statistical analysis paragraph 1 |
| | | (*d*) *Cohort study*—If applicable, explain how loss to follow-up was addressed | NA |
| | | (*e*) Describe any sensitivity analyses | Methods, Statistical analysis paragraph 3 |

| | | | |
|---|---|---|---|
| **Results** | | | |
| Participants | 13* | (a) Report numbers of individuals at each stage of study—e.g. numbers potentially eligible, examined for eligibility, confirmed eligible, included in the study, completing follow-up, and analysed | Methods, Study Population |
| | | (b) Give reasons for non-participation at each stage | Methods, Study Population |
| | | (c) Consider use of a flow diagram | Supporting Information, Figure S1 |
| Descriptive data | 14* | (a) Give characteristics of study participants (e.g. demographic, clinical, social) and information on exposures and potential confounders | Results, Participant Characteristics |
| | | (b) Indicate number of participants with missing data for each variable of interest | Supporting Information Figure S1 |
| | | (c) *Cohort study*—Summarise follow-up time (e.g., average and total amount) | NA |
| Outcome data | 15* | *Cohort study*—Report numbers of outcome events or summary measures over time | Results, Participant Characteristics, Pattern of multimorbidity |
| Main results | 16 | (*a*) Give unadjusted estimates and, if applicable, confounder-adjusted estimates and their precision (eg, 95% confidence interval). Make clear which confounders were adjusted for and why they were included | Results, Risk of severe SARS-CoV-2 infection |
| | | (*b*) Report category boundaries when continuous variables were categorized | Results, Risk of severe SARS-CoV-2 infection |
| | | (*c*) If relevant, consider translating estimates of relative risk into absolute risk for a meaningful time period | NA |
| Other analyses | 17 | Report other analyses done—e.g. analyses of subgroups and interactions, and sensitivity analyses | Results, Risk of severe SARS-CoV-2 infection, paragraph 2 |
| **Discussion** | | | |
| Key results | 18 | Summarise key results with reference to study objectives | Discussion paragraph 1 |
| Limitations | 19 | Discuss limitations of the study, taking into account sources of potential bias or imprecision. Discuss both direction and magnitude of any potential bias | Discussion paragraph 5 |
| Interpretation | 20 | Give a cautious overall interpretation of results considering objectives, limitations, multiplicity of analyses, results from similar studies, and other relevant evidence | Discussion paragraph 2-5 |
| Generalisability | 21 | Discuss the generalisability (external validity) of the study results | Discussion paragraph 6 |
| **Other information** | | | |
| Funding | 22 | Give the source of funding and the role of the funders for the present study and, if applicable, for the original study on which the present article is based. | End of the manuscript |
